# Relative burdens of the COVID-19, malaria, tuberculosis, and HIV/AIDS epidemics in sub-Saharan Africa

**DOI:** 10.1101/2021.03.27.21254483

**Authors:** David Bell, Kristian Schultz Hansen

**Author notes:** Corresponding author David Bell, Issaquah, WA 98027, USA.

## Abstract

COVID-19 has had considerable global impact, but in sub-Saharan Africa is one of several infectious disease priorities. Prioritization is normally guided by disease burden, but the highly age-dependent nature of COVID-19 and other infectious diseases makes comparisons challenging unless considered through metrics that incorporate life years lost and time lived in adverse health. We therefore compared 2020 mortality and Disability-Adjusted Life Years lost (DALYs lost) estimates for malaria, tuberculosis, and HIV/AIDS in sub-Saharan African populations with 12+ months of COVID-19 burden (up to end March 2021), applying known age-related mortality to United Nations estimates of age structure. We further compared exacerbations of disease burden predicted from the COVID-19 public health response. Data was derived from public sources, predicted exacerbations derived from those published by international agencies. For sub-Saharan African populations north of South Africa, recorded COVID-19 DALYs lost in 2020 was 3.7%, 2.3%, and 2.4% of those estimated for tuberculosis, HIV/AIDS and malaria respectively. Predicted exacerbations of these comparator diseases were greater than the estimated COVID-19 burden. Including South Africa and Lesotho, COVID-19 DALYs lost were <12% of those for comparator diseases and dominated by them in all age groups below 65 years. The analysis suggests a relatively low impact from COVID-19. While all four epidemics continue, tuberculosis, HIV/AIDS, and malaria remain far greater health priorities based on disease burden. Resource diversion to COVID-19 therefore runs a high risk of increasing the overall disease burden and causing net harm, further increasing global inequities in health and life expectancy.

## Introduction

COVID-19 has massively impacted life and society in sub-Saharan Africa, as elsewhere. Despite relatively low COVID-19 mortality rates in most countries of sub-Saharan Africa, aspects of the lockdown responses including business and school closures and restricted health service access introduced in the early days of the pandemic continue in various forms.^1^ In South Africa, infection with the virus itself has significantly impacted health, with 28469 deaths attributed by the end of 2020,^1^ but the younger aged populations to the north have recorded far lower mortality.^2^ Public health interventions must be tailored to address such variation. This requires realistic metrics for disease burden that take the characteristics of the population, and the individual impacted by disease, into account.^3^

Time-based measures such as Disability-Adjusted Life Years (DALYs) that incorporate loss of healthy life due to premature death and time lived in less-than-optimal health have potential to better represent the full impact of disease, as opposed to mortality alone. This is of particular relevance when the disease being measured is highly age-related. This is not a reflection on the value of human life – all lives are considered of equal value. The use of DALYs lost serves as an important guide in resource allocation to ensure that the greatest health impact is achieved. A young child dying from pneumonia is clearly expected to lose more potential life years than an 80-year-old dying from the same, so interventions based on metrics that prioritize childhood pneumonia will achieve greater overall impact. In resource-constrained sub-Saharan Africa, metrics of disease burden are of particular importance. Malaria imparts a disproportionate burden in disability adjusted life years (DALYs) lost, as most mortality occurs before age five. HIV/AIDS causes long spells of severe ill-health and premature death primarily among young and middle-aged adults, leading to significant life years lost and extended time lost due to less-than-optimal health.

COVID-19 is characterized by its strong association with advanced age, with a mean age of death similar to that for all-cause mortality in many countries.^5,6^ However, reporting of the burden of COVID-19 has generally ignored age of death, centering on comparisons of total mortality alone. Estimates based on total mortality also ignore the strong association of death from COVID-19 with pre-existing morbidities, which further reduce expected life years lost.^6^

Low COVID-19-associated mortality among sub-Saharan African populations is at least partially predicted by their young age structure,^5,6,7^ while lifestyle factors may be protective through a lower prevalence of major co-morbidities,^8-10^ higher vitamin D levels, and broad antigen exposure leading to non-specific T-cell prior immunity.^11-13^ Comparisons of lockdown severity suggest that more restrictive measures have had limited additional impact on reducing COVID-19 mortality.^14-16^ However, as with many public health responses, lockdown responses are not without cost. Predicted exacerbations of high burden diseases including malaria, HIV/AIDS, and tuberculosis impact particularly children and younger adults.^17-19^ Broader impacts of reduced food security and interruption of vaccination will have far-reaching health consequences,^20,21^ while loss of family income and reductions in national gross domestic product will impede the capacity to respond.^22^

While lockdowns may be easing, proposals for continent-wide mass vaccination under the COVAX mechanism will raise new costs and divert resources, and the urgency of developing good public health policy that appropriately prioritizes management of diseases including COVID-19, based on their relative burdens, is no less urgent.^23^ We therefore compared the disease burdens of COVID-19 and the three pre-existing major infectious disease ‘epidemics’ of sub-Saharan African countries,^24^ with and without South Africa and Lesotho, to estimate the relative burden of COVID-19 in relation to these other epidemics.

## Methods

### Health indicators and geography

The health indicators used for the analyses were the number of deaths and Disability-Adjusted Life Years (DALYs) lost by age group caused by COVID-19 and three major diseases: malaria, HIV/AIDS, and tuberculosis. Published data were sourced for the analyses. For the calculation of DALYs lost caused by these diseases at population level, the estimates for number of deaths by age and the number of non-fatal episodes of ill-health by age in 2020 were required. The geographical area assessed, ‘sub-Saharan Africa’, excludes the five countries bordering the Mediterranean Sea. Estimates with and without South Africa and Lesotho were included as these two countries have very different burdens of the four diseases, and differing demographics.

### COVID-19 mortality and DALYs lost

As COVID-19 cases were reported from Africa in February 2020 and widely spread by late March, all recorded COVID-19 deaths were included up until 31 March 2021 to include slightly over 12 months of reporting. According to Africa CDC there had been 77,463 reported COVID-19 deaths in sub-Saharan Africa up to March 2021 and 24,302 COVID-19 deaths if South Africa and Lesotho were excluded.^2^ These total reported COVID-19 deaths were allocated across age groups and also used to estimate the number of COVID-19 infections by applying the following method and assumptions. Using data on COVID-19 deaths and seroprevalence surveys from 45 mainly European countries, O’Driscoll et al. estimated the infection fatality ratio by age.^25^ Assuming that these estimated infection fatality ratios also represented the sub-Saharan African situation and further assuming a constant share of infection across age groups, the number of infections and deaths were inferred for a sub-Saharan Africa population size according to United Nations population estimate for 2020 and compatible with a total number of 77,463 (24,302) COVID-19 deaths (see Table S1 in the supplementary material).^26^ These estimated numbers of deaths and infections by age group were inserted in the standard DALY formula used for calculating the burden of disease with no discounting of future life years and without the age weighting function.^27^ For the calculation of life years lost, the standard life expectancies by age from the Global Burden of Disease Study 2019 were used,^28^ and the reference life table was downloaded from the Institute of Health Metrics and Evaluation website.^29^ The non-fatal COVID-19 infections were assumed to be mild, of a two-week duration, and a disability weight of 0.051 was used, corresponding to the weight attached to a moderate to severe upper respiratory infection.^28^

### Mortality and DALYs lost for HIV/AIDS, tuberculosis, and malaria

The number of deaths and DALYs lost for HIV/AIDS and tuberculosis by age group for 2019 were extracted from the Global Burden of Disease Study 2019 results.^29^ These estimates were updated to 2020 by assuming a growth from 2019 to 2020 corresponding to the annual population growth rate in sub-Saharan Africa. Population growth rates were estimated by age group using population estimates from 2015 and 2020.^26^ Cases of combined HIV/AIDS and tuberculosis are considered as HIV/AIDS only and not included in the tuberculosis burden calculations.

The total number of deaths and non-fatal illness episodes caused by malaria in sub-Saharan Africa were obtained from the World Health Organization (WHO) estimates for 2019.^4^ However, the published numbers were not available by age group. It was assumed that the deaths and illness spells followed the same distribution across age groups as malaria deaths estimated by the Institute of Health Metrics and Evaluation.^29^ The number of malaria deaths and illness episodes by age in sub-Saharan Africa (with and without South Africa and Lesotho) were translated into DALYs lost applying the same method as described above including the assumption that a non-fatal malaria infection lasted two weeks and with a disability weight of 0.051.

### Predicted exacerbations of HIV/AIDS, tuberculosis and malaria

Predicted exacerbations of the three comparator diseases accrued from 2020 lockdown responses were derived from modelling published by WHO, StopTB Partnership and The Global Fund, simulating effects on transmission of reduced healthcare access and, for malaria, reduced vector control.^17-19^

### Patient and Public Involvement

There was no patient involvement in this study. All data was obtained from publicly available sources.

### Ethics approval

The analysis is based entirely on publicly available data, and no specific ethical approval was required.

## Results

Recorded COVID-19 mortality constituted 6.4%, 4.8%, and 6.3% of the mortality of tuberculosis, HIV/AIDS, and malaria respectively in sub-Saharan Africa north of South Africa and Lesotho. As DALYs lost, the COVID-19 burden amounted to 3.7%, 2.3%, and 2.4% of that estimated for the three comparator diseases respectively (Figure 1). Including South Africa and Lesotho, recorded COVID-19 mortality was 19.2%, 11.7%, and 20.2% of the mortality of tuberculosis, HIV/AIDS, and malaria respectively, while as DALYs lost the COVID-19 burden amounted to 11.1%, 5.5%, and 7.5% respectively of DALYs lost for these three comparator diseases (Figure 1).

**Figure 1.**
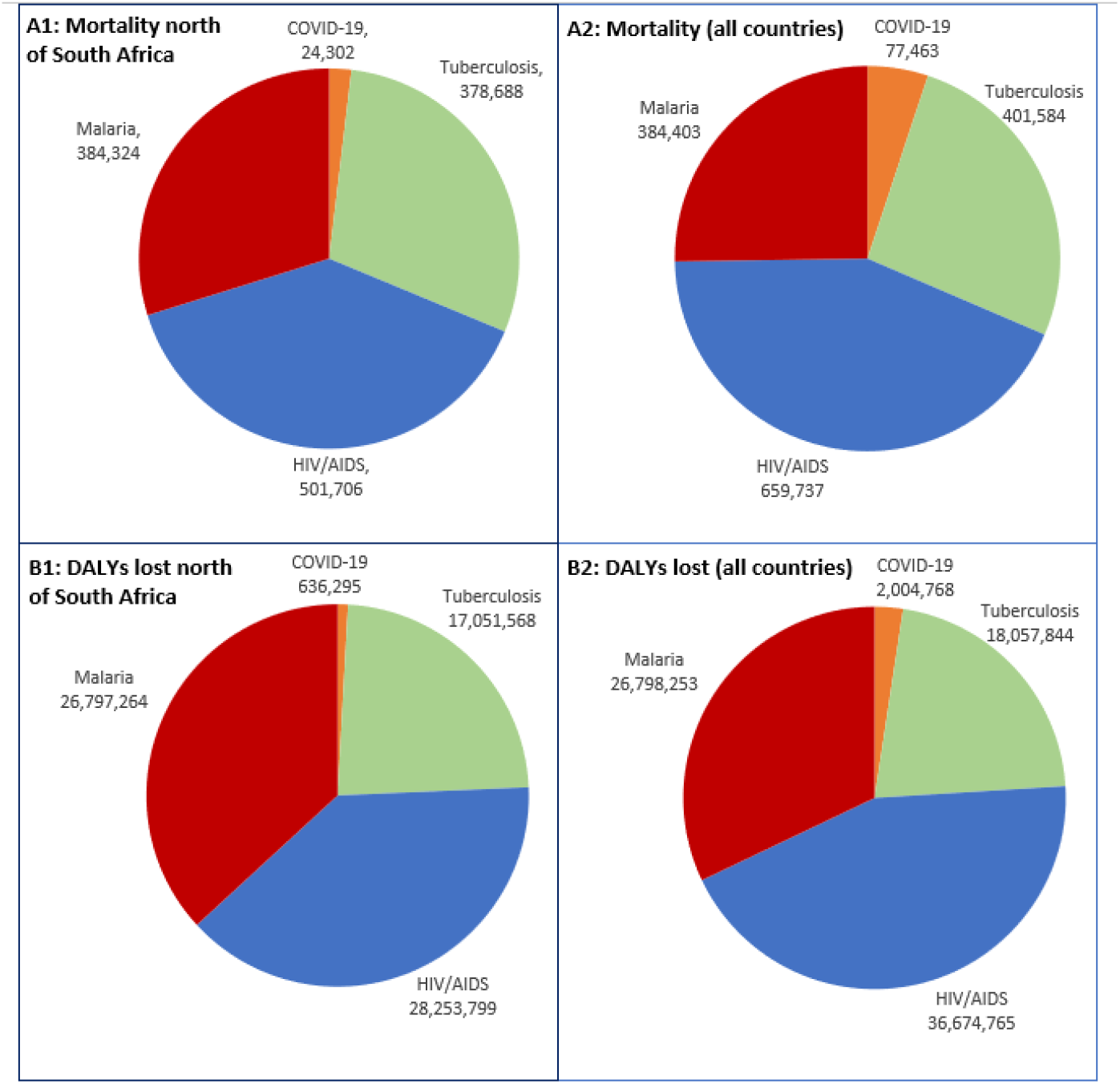
Comparison of baseline mortality and disease burdens (DALYs lost) predicted for the 12 months of 2020 for malaria, tuberculosis, and HIV/AIDs (pre-lockdown impact), and up to 31 March 2021 for COVID-19, in sub-Saharan Africa. A1 Mortality for sub-Saharan countries north of South Africa and Lesotho. A2: Mortality for all sub-Saharan countries. B1 DALYs lost for sub-Saharan countries north of South Africa and Lesotho. B2: DALYs lost for all sub-Saharan countries.

Tuberculosis mortality was never dominated by COVID-19 in any age-group, while HIV/AIDS was only dominated above 70-74 years of age and malaria dominated above 75-79 years in sub-Saharan Africa north of South Africa and Lesotho (Figure 2). Including South Africa and Lesotho, all three comparator diseases individually dominated COVID-19 mortality until 65-69 years, after which COVID-19 dominated HIV/AIDS, while malaria was also dominated after 70-74 years (Figure 2).

**Figure 2.**
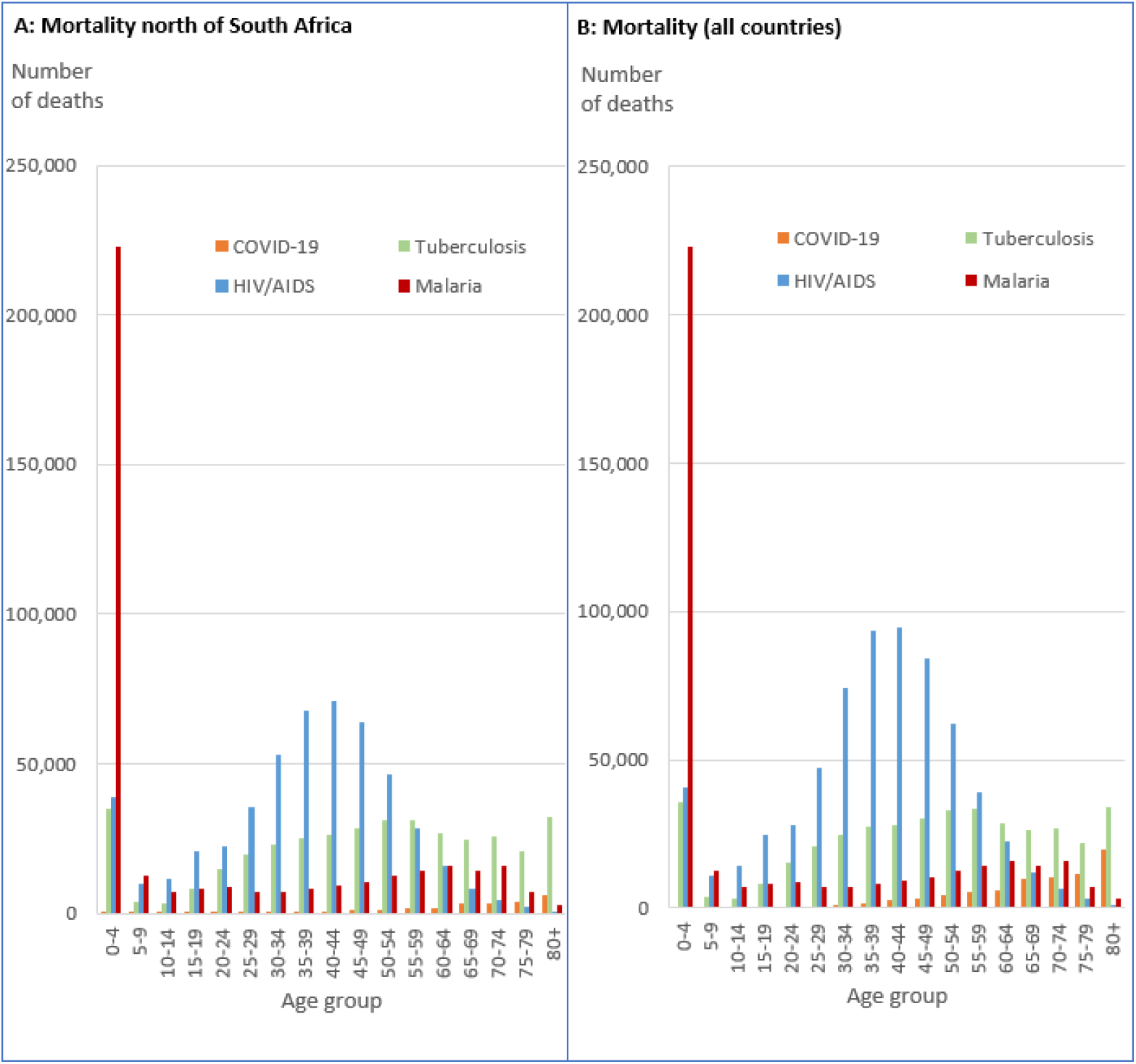
Mortality predicted for the 12 months of 2020 for malaria, tuberculosis, and HIV/AIDs (pre-lockdown impact), and up to 31 March 2021 for COVID-19, in sub-Saharan Africa. A: Mortality for sub-Saharan countries north of South Africa and Lesotho. B: Mortality for all sub-Saharan countries.

Accrued COVID-19 DALYs lost up to 31 March 2021 were lower than the lower-limit estimates of exacerbation of malaria, tuberculosis, and HIV/AIDS (increase over baseline) predicted due to the COVID-19 public health response in sub-Saharan Africa north of South Africa, and below those for tuberculosis and HIV/AIDS for sub-Saharan Africa as a whole. If we assume significant under-reporting of COVID-19 mortality and arbitrarily multiply by a factor of 10, DALYs lost from COVID-19 only dominates the lower predicted exacerbations of malaria-burden in sub-Saharan Africa north of South Africa. It remains below all baseline disease burdens for malaria and HIV/AIDs in sub-Saharan Africa as a whole, and all comparator diseases where South Africa and Lesotho are excluded (Table 1).

**Table 1.**
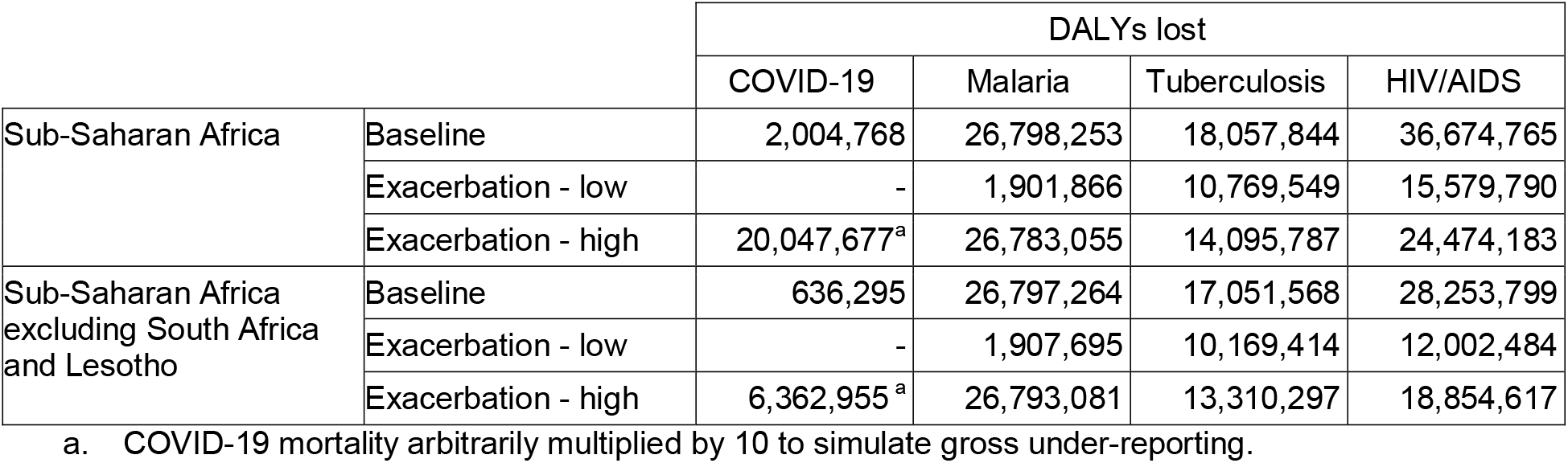
Comparisons of recorded disease burden (DALYs lost) from COVID-19 in sub-Saharan Africa up till 31 March 2021 compared to predicted exacerbations for malaria, tuberculosis, and HIV/AIDS from the impact of the COVID-19 public health response.

|  |  | DALYs lost |  |  |  |
| --- | --- | --- | --- | --- | --- |
|  |  | COVID-19 | Malaria | Tuberculosis | HIV/AIDS |
| Sub-Saharan Africa | Baseline | 2,004,768 | 26,798,253 | 18,057,844 | 36,674,765 |
|  | Exacerbation - low | - | 1,901,866 | 10,769,549 | 15,579,790 |
|  | Exacerbation - high | 20,047,677<sup>a</sup> | 26,783,055 | 14,095,787 | 24,474,183 |
| Sub-Saharan Africa<br>excluding South Africa<br>and Lesotho | Baseline | 636,295 | 26,797,264 | 17,051,568 | 28,253,799 |
|  | Exacerbation - low | - | 1,907,695 | 10,169,414 | 12,002,484 |
|  | Exacerbation - high | 6,362,955<sup>a</sup> | 26,793,081 | 13,310,297 | 18,854,617 |

a. COVID-19 mortality arbitrarily multiplied by 10 to simulate gross under-reporting.

## Discussion

These comparisons emphasize the relatively low disease burden that COVID-19 is exerting on sub-Saharan African populations, with the pre-existing ‘epidemics’ of malaria, tuberculosis, and HIV/AIDS all greatly dominating in life-years impacted, with mortality dominating also in all except the elderly. The use of DALYs lost to assess relative disease burden follows well-accepted practice,^30,31^ but is relatively unusual for assessment of COVID-19. It is unclear why this standard public health metric, or the related metric of Quality-Adjusted Life Years (QALYs), has not been widely applied to a disease with such a characteristic age-dependent impact. Appropriate assessment of burden is of extreme importance as decisions are made globally on resources to be allocated to vaccination as well as the imposition of recurrent lockdowns and other cost-bearing responses. Mass vaccination of the sub-Saharan African population against COVID-19, as is advocated in some quarters,^23^ will draw essential resources from interventions aimed at health problems with far greater burden through diversion of financial resources and personnel on the ground. Such a wide-scale vaccination intervention has never been attempted before and the implications for already over-stretched health services will be significant. To ensure equity in health care, a comprehensive economic evaluation comparing costs and effects of interventions against all four epidemics, including cost-effectiveness analysis, is urgently needed.

This data analysis has a number of limitations. COVID-19 mortality reporting in sub-Saharan Africa is doubtless incomplete, though low mortality is predicted by population age structure and lower prevalence of major co-morbidities including obesity,^5,7,32^ while other lifestyle factors and prior immunity may also be protective.^10,12,33,34^ Given the lack of strong local data on age-related mortality, we assumed that this will reflect rates found elsewhere.^25^ Lack of transmission appears an unlikely explanation for low recorded mortality as high seroprevalence has been recorded in various sub-Saharan African settings.^33-39^ While the higher mortality of COVID-19 in South Africa could be partially explained by higher reporting rates, South Africa also has higher rates of known mortality risk factors.^40^ Evidence of very high asymptomatic infection,^39^ and the level of testing taking place (868,823 tests for 333 deaths in Uganda alone by 23 February 2021),^41^ suggests that the relatively low recorded mortality in most countries reflects reality, in common with much of Asia.^42^

The relative burden of COVID-19 in 2020 is also subject to the first cases only being reported in March in most of these populations,^42^ and so it may not have spread to all populations within the first few months of recording. However, the total mortality rate remains low across the continent at time of writing.^42^ While significant increases have occurred in countries bordering South Africa from mid-2021, reported mortality in the more populous countries to the north has remained low by global comparisons.^42^ The bulk of the COVID-19 burden may well been accrued in the >12 months total recording time used here.

DALYs lost through COVID-19 morbidity as estimated in this paper do not take post-viral syndromes into account (e.g., ‘long-covid’). These have limited prevalence and may be lighter in younger (less severe illness) African populations,^43^ but this is still unclear and will add somewhat to COVID-19 burden. Conversely, the age-based nature of DALYs lost applied to COVID-19 does not take into account the high prevalence of life-shortening comorbidities associated with these cases,^6^ which will in turn lead to an over-estimation of the actual life years lost. Even assuming 90% under-estimation of COVID-19 mortality here, malaria and HIV/AIDS disease burdens still dominate COVID-19, as do most upper estimates of exacerbation of these through the COVID-19 public health response.

In comparing impact of COVID-19 and other health burdens, we considered just three diseases. The broad impact of malnutrition, and of reduced educational attainment (closed schools) and damage to local and national economies will have major long-term impacts on population and societal health.^20-22,44^ As a greater proportion of the population achieves post-infection immunity,^35-37,41,45,46^ COVID-19 burden is likely to further reduce, and the cost-effectiveness of response interventions may then decrease further. It is therefore imperative that cost-effectiveness analyses of further public health responses to COVID-19 in these sub-Saharan African populations be tailored to local need, based on realistic metrics that reflect the impact of COVID-19 relative to other high-burden diseases. These analyses should also account for any potential negative impacts of these responses, including reduced health system access and resource diversion that may result from severe lockdowns or mass vaccination. Failure to address this will risk increasing health inequities rather than reducing them.

## Data Availability

All data is in the public domain, and calculations are freely available from the authors.

## List of abbreviations

Africa CDC: Africa Centres for Disease Control and Prevention
AIDS: Acquired Immune Deficiency Syndrome
DALYs: Disability-Adjusted Life Years
HIV: Human Immunodeficiency Virus
SARS-CoV-2: Severe Acute Respiratory Syndrome Corona Virus -2
WHO: World Health Organization

## Availability of data

All data is in the public domain, and calculations are freely available from the authors.

## Author contributions

DB and KSH jointly conceived the study, contributed to the study design and conceptualization, and sourced the data. KSH performed the economic analyses, both authors participated in the drafting, revising and approval of the manuscript.

## Patient and public involvement

There was no direct patient or public involvement in this study. All data used was in the public domain.

## SUPPLEMENTAL FILE

## Supplementary information

**Table S1a.** Estimated COVID-19 infections (individuals infected) and deaths by age in Sub-Saharan Africa excluding South Africa and Lesotho, up to 31 March 2021.

| Age group | United Nations<br>population<br>estimate 2020<sup>a</sup> | Share of<br>population<br>infected (%)<sup>b</sup> | Infection<br>fatality<br>ratio (%)<sup>c</sup> | Estimated<br>COVID-19<br>infections | Estimated<br>COVID-19<br>deaths |
| --- | --- | --- | --- | --- | --- |
| 0-4 | 164,804,603 | 2.282 | 0.003 | 3,761,278 | 113 |
| 5-9 | 147,530,309 | 2.282 | 0.001 | 3,367,033 | 34 |
| 10-14 | 130,237,158 | 2.282 | 0.001 | 2,972,357 | 30 |
| 15-19 | 112,354,975 | 2.282 | 0.003 | 2,564,238 | 77 |
| 20-24 | 95,039,505 | 2.282 | 0.006 | 2,169,053 | 130 |
| 25-29 | 79,611,837 | 2.282 | 0.013 | 1,816,953 | 236 |
| 30-34 | 66,961,607 | 2.282 | 0.024 | 1,528,241 | 367 |
| 35-39 | 55,972,752 | 2.282 | 0.040 | 1,277,447 | 511 |
| 40-44 | 45,533,164 | 2.282 | 0.075 | 1,039,187 | 779 |
| 45-49 | 36,505,391 | 2.282 | 0.121 | 833,150 | 1,008 |
| 50-54 | 28,697,760 | 2.282 | 0.207 | 654,959 | 1,356 |
| 55-59 | 22,713,657 | 2.282 | 0.323 | 518,386 | 1,674 |
| 60-64 | 17,441,001 | 2.282 | 0.456 | 398,050 | 1,815 |
| 65-69 | 12,682,251 | 2.282 | 1.075 | 289,443 | 3,112 |
| 70-74 | 8,585,543 | 2.282 | 1.674 | 195,945 | 3,280 |
| 75-79 | 5,010,511 | 2.282 | 3.203 | 114,353 | 3,663 |
| 80+ | 3,232,639 | 2.282 | 8.292 | 73,777 | 6,118 |
| Total | 1,032,914,663 |  |  | 23,573,850 | 24,302 |

<sup>a</sup> Reference (7).
<sup>b</sup> Assumed population share compatible with 13,031 COVID-19 deaths in total.
<sup>c</sup> Reference (25).

**Table S1b.**
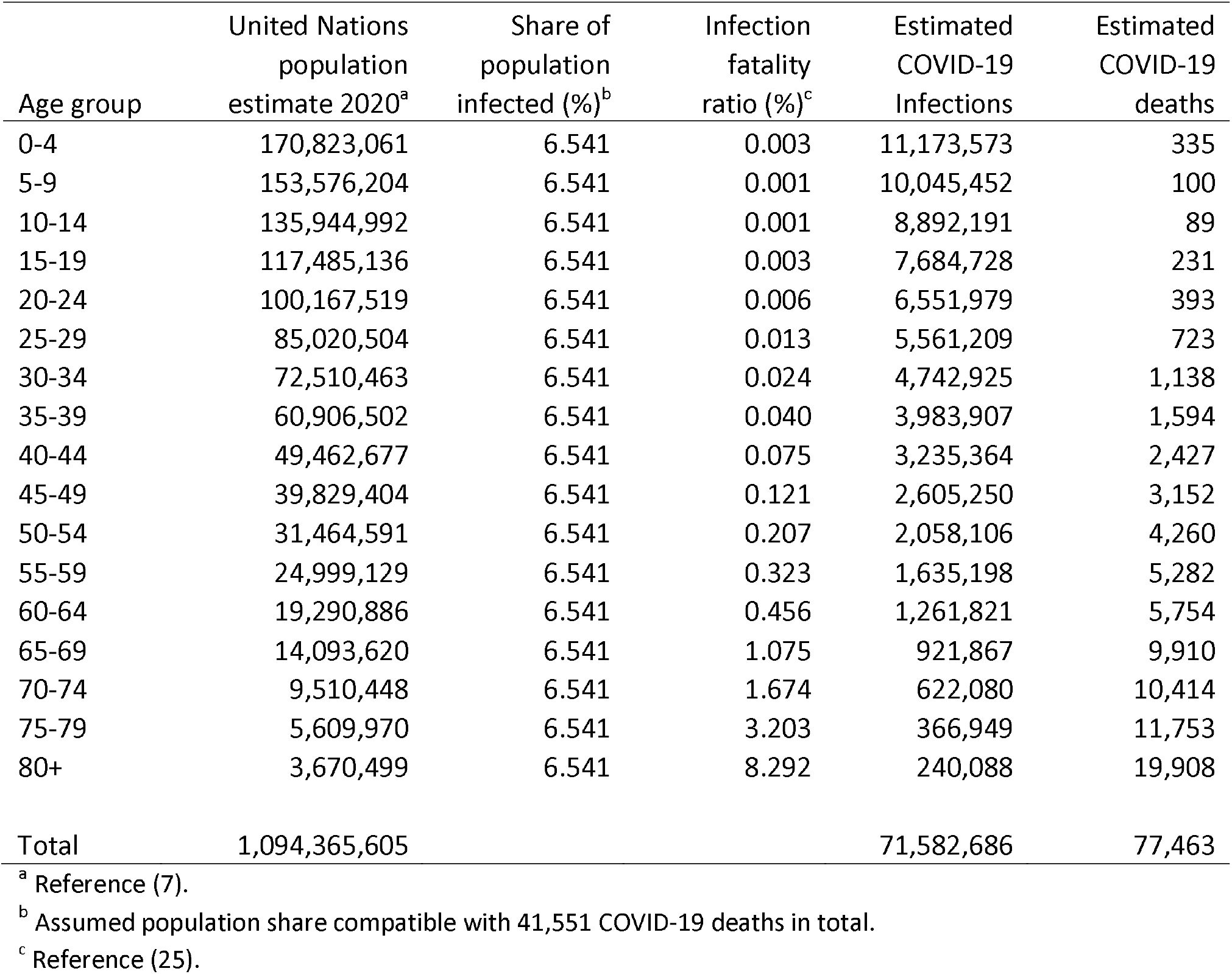
Estimated COVID-19 infections (individuals infected) and deaths by age in Sub-Saharan Africa, up to 31 March 2021.

